# A novel molecular analysis approach in colorectal cancer suggests new treatment opportunities

**DOI:** 10.1101/2023.01.13.23284526

**Authors:** Elena López-Camacho, Guillermo Prado-Vázquez, Daniel Martínez-Pérez, María Ferrer-Gómez, Sara Llorente-Armijo, Rocío López- Vacas, Mariana Díaz- Almirón, Angelo Gámez-Pozo, Juan Ángel Fresno Vara, Jaime Feliu, Lucía Trilla-Fuertes

**Author notes:** Corresponding author: Lucía Trilla-Fuertes, Molecular Oncology Lab, La Paz University Hospital-IdiPAZ, Paseo de la Castellana 261, 28046, Madrid, Spain.

## Abstract

Colorectal cancer (CRC) is a molecular and clinically heterogeneous disease. In 2015, the Colorectal Cancer Subtyping Consortium classified CRC into four consensus molecular subtypes (CMS), but these CMS have had little impact on the clinical practice. The purpose of this study is to deepen into the molecular characterization of CRC. A novel approach, based on probabilistic graphical models (PGM) and sparse k-means-Consensus Cluster layer analyses was applied in order to functionally characterize CRC tumors. First, PGM was used to functionally characterize CRC, and then, sparse k-means-Consensus cluster was used to explore layers of biological information and establish classifications. To this aim, gene expression and clinical data of 805 CRC samples from three databases were analyzed. Tree different layers based on biological features were identified: adhesion, immune and molecular. The adhesion layer divided patients into high and low adhesion groups, with prognostic value. The immune layer divided patients into immune-high and immune-low groups, according to the expression of immune-related genes. The molecular layer established four molecular groups related to stem cells, metabolism, Wnt signalling pathway and extracellular functions. Immune-high patients, with a higher expression of immune-related genes and genes involved in viral mimicry response may be benefit for immunotherapy and viral mimicry-related therapies. Additionally, several possible therapeutic targets have been identified in each molecular group. Therefore, this improved CRC classification could be useful for searching new therapeutic targets and specific therapeutic strategies in CRC disease.

**Simple summary:** Colorectal cancer is a heterogeneous disease. Several efforts have been done to characterize this heterogeneity but they have not impact in clinic. In this work, we used a novel analysis approach based on identifying layers of information using expression data from colorectal tumors and characterize three different layers of information: one layer related to adhesion with prognostic value, one related to immune characteristics and one related to molecular features. The molecular layer divided colorectal tumors in stem cell, Wnt, metabolic, and extracellular groups. These molecular groups suggested some possible therapeutic targets for each group. Additionally, immune characteristics divided tumors in tumors with a high expression of immune and viral mimicry response genes and with a low expression, suggesting immunotherapy and viral mimicry related therapies as suitable for these immune-high patients.

## 1. Introduction

Colorectal cancer (CRC) has been identified as the most prevalent tumor. According to GLOBOCAN, there were 19.3 million of new cases and 10 million of cancer deaths worldwide in 2020. CRC ranks third in terms of incidence, representing the 10 % of new cancer cases, but the second in terms of mortality, with 940,000 estimated deaths [1]. CRC is a molecularly heterogenic disease, in which molecular alterations influence the growth and survival of tumor cells, as well as their differentiation, apoptosis, and distant metastasis [2]. The heterogeneity presented by this cancer has also been related to the anatomical location of the tumor, since the proximal and distal colon have different embryological origins [3]. In this context, Bufill *et al*. established the first classification of colorectal cancer, defining two groups: group I or proximal if the tumor was located on the right side, and group II or distal when located on the left side [4]. The American Joint Committee (AJCC) on Cancer tumor-node-metastasis (TNM) staging system is the most common staging system in clinical settings. However, a detailed analysis of the prognostic significance of the 8^th^ edition TNM classification for CRC tumors showed that this staging system is not enough accurate for evaluating the prognosis of CRC in the clinic [5].

Until 2015, different genetic classifications for CRC have been proposed. In that year, the international CRC Subtyping Consortium (CRCSC) was created a consensus on molecular genetic expression subtyping of CRC using a pooled molecular genetic analysis of 4151 colon tumors. Four colon cancer consensus molecular subtypes (CMS) were identified: CMS1 (microsatellite instability immune, 14%), CMS2 (canonical, 37%), CMS3 (metabolic, 13%), and CMS4 (mesenchymal, 23%), 13% of the samples could not be classified into any of the four described molecular subtypes [6]. These unclassified tumors could present high intratumor heterogeneity or correspond to an intermediate phenotype, with characteristics belonging to different molecular subtypes [7]. In non-metastatic disease, the poor prognostic value of CMS4 and the relatively favorable prognosis of CMS1 and CMS2 have been established [7]. Moreover, different studies established associations of CMS with treatment outcomes [8-10], and their potential for clinical use in predicting both prognosis and response to systemic therapy has been recently evaluated with encouraging results [6]. Clinical and therapeutic utility of the different molecular classifications has been discussed [11] but, despite the increasing knowledge, treatments based on a molecular subtype are not currently used in the clinical decision making [12].

Computational analyses applied to high-dimensional omics data allow a deeper characterization of the molecular and immune features of tumors. Probabilistic graphical models (PGMs) have been previously used to identify differences in biological processes among several tumors types [13-18]. Classification methods, such as sparse k-means [19], and Consensus Cluster (CC) [20], have previously demonstrated their utility in the establishment of tumor and immune subtypes for breast and bladder cancers [13, 18].

The main objective of this study is to expand the knowledge about the molecular classification of CRC, according to the different biological realities of the tumor, with the aim to increase the clinical value of the already established molecular groups.

## 2. Material and methods

### 2.1. Data search and curation

Three colorectal tumor gene expression databases (GSE17536, GSE35896, and GSE39582) were analyzed. The resulting database was processed, removing control and duplicated probes. For this purpose, the variance of each probe was calculated and the most variable probe per gene was chosen. In addition, the batch effect due to the combination of independent databases was corrected using the *limma* R package [21]. Information about the CMS group of each sample was downloaded from the Synapse platform [22]. Finally, clinical data from the three databases were collected and unified for further analysis.

### 2.2 Gene selection and probabilistic graphical model analysis

First, those genes with the higher standard deviation in their expression across the dataset (standard deviation >ill2) were selected to build the PGM, as previously described [18]. Briefly, gene expression data were used without other a priori information and the analyses were done using *grapHD* package [23] and R v3.2.5 [24]. PGMs are undirected acyclic graphs built in two steps: in the first step, the spanning tree that maximizes the likelihood was found, and then, the graph which preserved the decomposability and minimizing the Bayesian Information Criterion (BIC) with the simplest structure was chosen by a forward search that successively adding edges. The resulting network was split into several branches and the most representative function of each branch was established by gene ontology analyses using DAVID 6.8 webtool [25]. “*Homo sapiens*” was used as background and categories Biocarta, GO-FAT and KEGG were selected.

To make comparisons between groups of samples, functional node activities were calculated as previously described [18]. Briefly, the mean expression of all the genes included in one branch related to the main function of this branch was calculated. Differences in functional node activity were assessed by non-parametric tests.

### 2.3. Biological layer analyses

Sparse K-means and Consensus cluster algorithm (CC) were used to explore the molecular information of CRC samples, as previously described [26]. Sparse K-means assigns a weight to each gene according to its relevance explaining the main variability source on the database. Then, using the genes that were selected by sparse k-means, CC was applied to define the optimum number of groups for each case. Once genes relevant to a layer of information were identified, they were removed from the dataset and the analysis was done again with the remaining genes, allowing the identification of different layers of information. Once the information layers were generated, gene ontology analyses were performed for each layer to derive functional information. Sparse k-means was performed using *sparcl* package [19] and CC was performed using *Consensus Cluster Plus* package [20] and R v3.2.5 [24].

Then, we used the biological layer information to establish different classifications based on different tumor features. Differential expression patterns among groups were analyzed by Significance Analysis of Microarrays (SAM), defining a false discovery rate (FDR) below 5% [27]. These analyses were carried out using TM4 Multiexperiment Viewer (MeV) 4.9 software [28].

### 2.4. Statistical analyses

GraphPad Prism v6 was used for basic statistical analyses. Network visualization were done in Cytoscape software [29]. Differences in node activity were evaluated using the Kruskal-Wallis comparison method and Dunn’s multiple comparison tests. Survival curves were estimated using the Kaplan-Meier method and compared with the log-rank test, using disease-free survival (DFS) as the event. DFS was defined as the time elapsed between surgery and new onset of disease. All p-values were two-sided and considered statistically significant below 0.05.

## 3. Results

### 3.1. Pre-processing of gene expression and clinical data

Gene expression datasets (GSE17536, GSE17536 and GSE39582) from the Gene expression omnibus repository [30] were merged, including probes with expression data in all datasets. After duplicated probes were removed, batch effect was corrected and a variability filter was applied, dataset with 1,700 genes and 805 samples was obtained. 177 samples were from the GSE17536, 62 samples from GSE35896, and 566 samples from GSE39582.

### 3.2. Patient characteristics

RNA-seq data from eight hundred and five CRC patients was used in this study. Clinical characteristics of the cohort were summarized in Sup Table 1. The median of follow-up was 37 months and 36 relapses occurred.

132 samples were assigned to CMS1 (16.3%), 315 samples were assigned to CMS2 (39%), 97 samples were assigned to CMS3 (12%) and 188 samples were assigned to CMS4 (23.3%). Thus, 73 samples (9.4%) were not assigned to any CMS.

### 3.3. Functional characterization

A PGM was built with the gene expression profile of the 1,700 more variable genes. Seeking for functional structure, 13 functional nodes with an overrepresented biological function were defined: immune, adhesion, inflammatory response, somatic stem cell, extracellular matrix, cellular response, extracellular response, nucleus, Wnt signaling pathways, plasmatic membrane, regulation of cardiac conduction, transport, and metabolism (figure 1 and supplementary table 2)).

**Figure 1:**
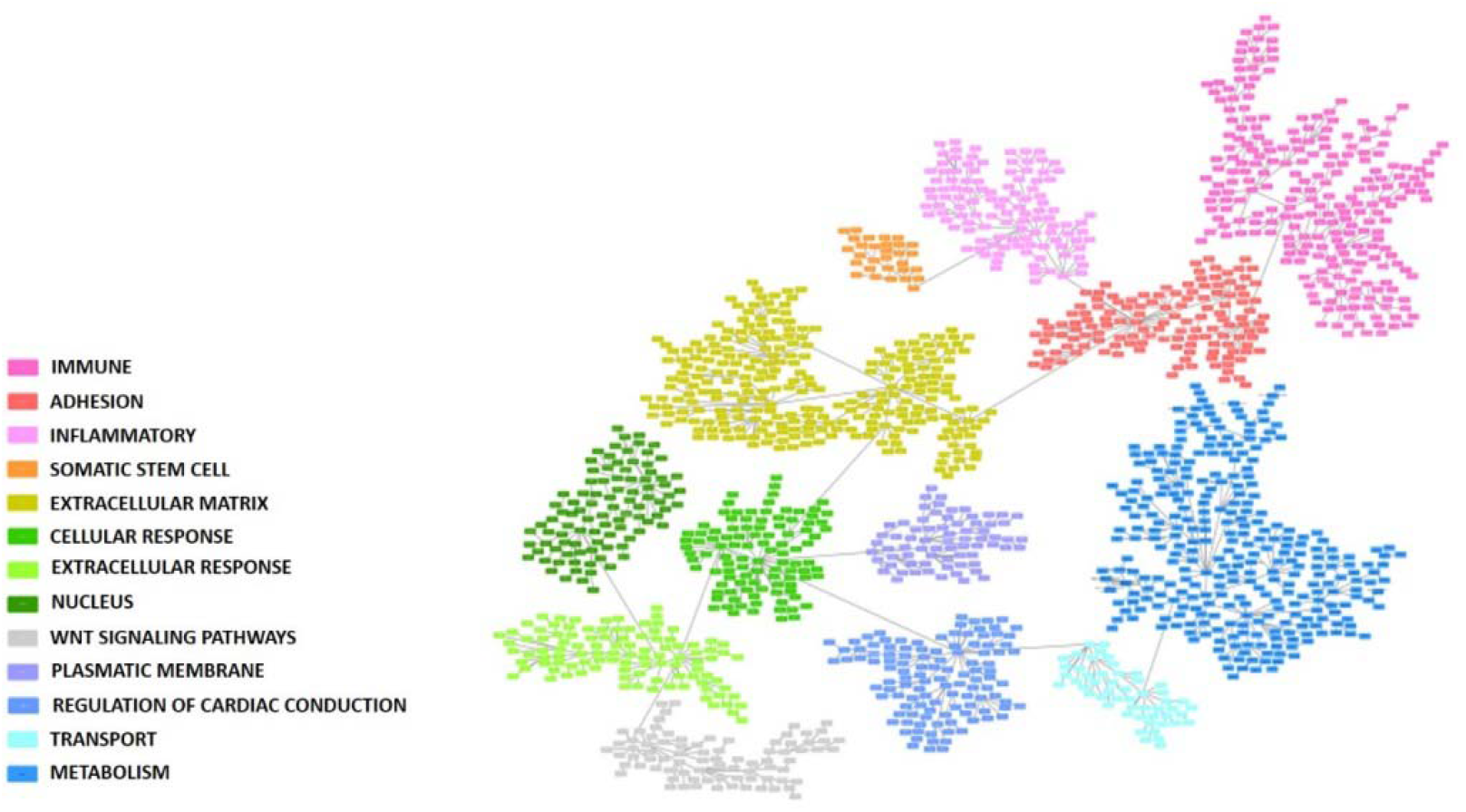
Probabilistic graphical model built using the 1,700 more variable genes in the colorectal cancer cohort. Each box represents one gene. Functional nodes are highlighted in the PGM.

### 3.4. Biological layer analysis

The sparse k-means-CC workflow analyses identified nine biological layers. Each layer was split into two groups with the exception of the second one, which optimal classification turned out to be in three groups. Each layer main function was characterized through gene ontology (Table 1). Layer 1 and 5 were related to adhesion processes, layers 2 and 6 were related to metabolic pathways and layers 3 and 8 were related to immune response. The two adhesion layers and the two immune ones were equivalent, respectively, dividing CRC patients into similar groups (Sup Figure 1). Therefore, three main biological layers of information were established: an adhesion layer, an immune layer and a molecular layer, the last one grouping all the information provided by metabolic, extracellular, and digestion classifications.

**Table 1:** Biological molecular layers obtained by the sparse k-means-consensus cluster workflow analyses and the number of groups in which CRC patients were divided according to each layer.

| Layers | Genes | Number of groups | Main gene ontology |
| --- | --- | --- | --- |
| 1<sup>st</sup> | 98 | 2 | Cellular adhesion |
| 2<sup>nd</sup> | 53 | 3 | Metabolic pathways |
| 3<sup>rd</sup> | 131 | 2 | Immune response |
| 4<sup>th</sup> | 32 | 2 | Digestion |
| 5<sup>th</sup> | 148 | 2 | Cellular adhesion |
| 6<sup>th</sup> | 92 | 2 | Metabolic pathways |
| 7<sup>th</sup> | 78 | 2 | Extracellular response |
| 8<sup>th</sup> | 89 | 2 | Inflammatory response |
| 9<sup>th</sup> | 88 | 2 | Ion calcium and cellular transport |

### 3.5. Adhesion layer

First and fifth layers were related to cellular adhesion, and divided the samples in a redundant way across the database, so both layers were merged into the Adhesion layer. Genes included in Adhesion layer were showed on Supplementary Table 3. CC determined that samples should be divided by their adhesion features into two different groups: adhesion 1, including 454 samples, with lower expression of the adhesion genes, and adhesion 2, including 351 samples, with higher expression of the adhesion genes (Figure 2A). Differentially expressed genes between Adhesion groups were identified using SAM and their mainly codified for proteins located in the extracellular matrix, like collagens, and were related with adhesion functions (figure 2B and supplementary table 4). Patients with low adhesion tumors had better prognosis (p= 0.0098, HR= 0.42, 95%CI=(0.21-0.81)) (Figure 3). Additionally, there were differences in DFS combining the information of the tumor stage and adhesion groups, having adhesion-high stage 3/4 tumors significantly worse prognosis and being adhesion-low stages3/4 tumors comparable in prognosis to adhesion-high stages 1/2 tumors (Figure 3B). No differences in the distribution in each adhesion group according to tumor location and stage were found.

**Figure 2:**
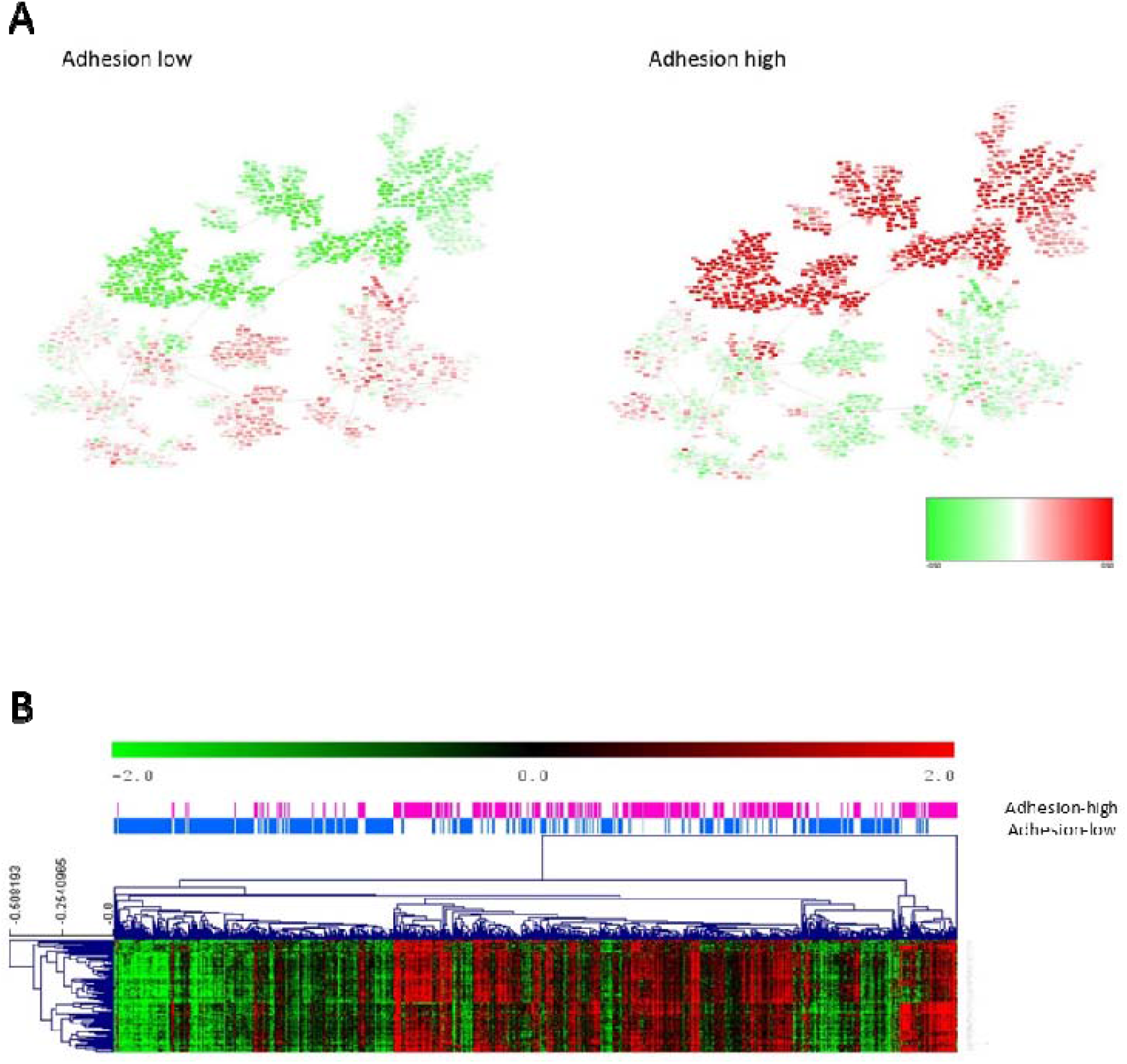
**A**. Heatmap of the mean gene expression of adhesion groups in the PGM. Red=overexpressed. Green= underexpressed. **B**. Differential expressed 100 genes in the adhesion layer between adhesion 1 and 2 groups identified by SAM.

**Figure 3:**
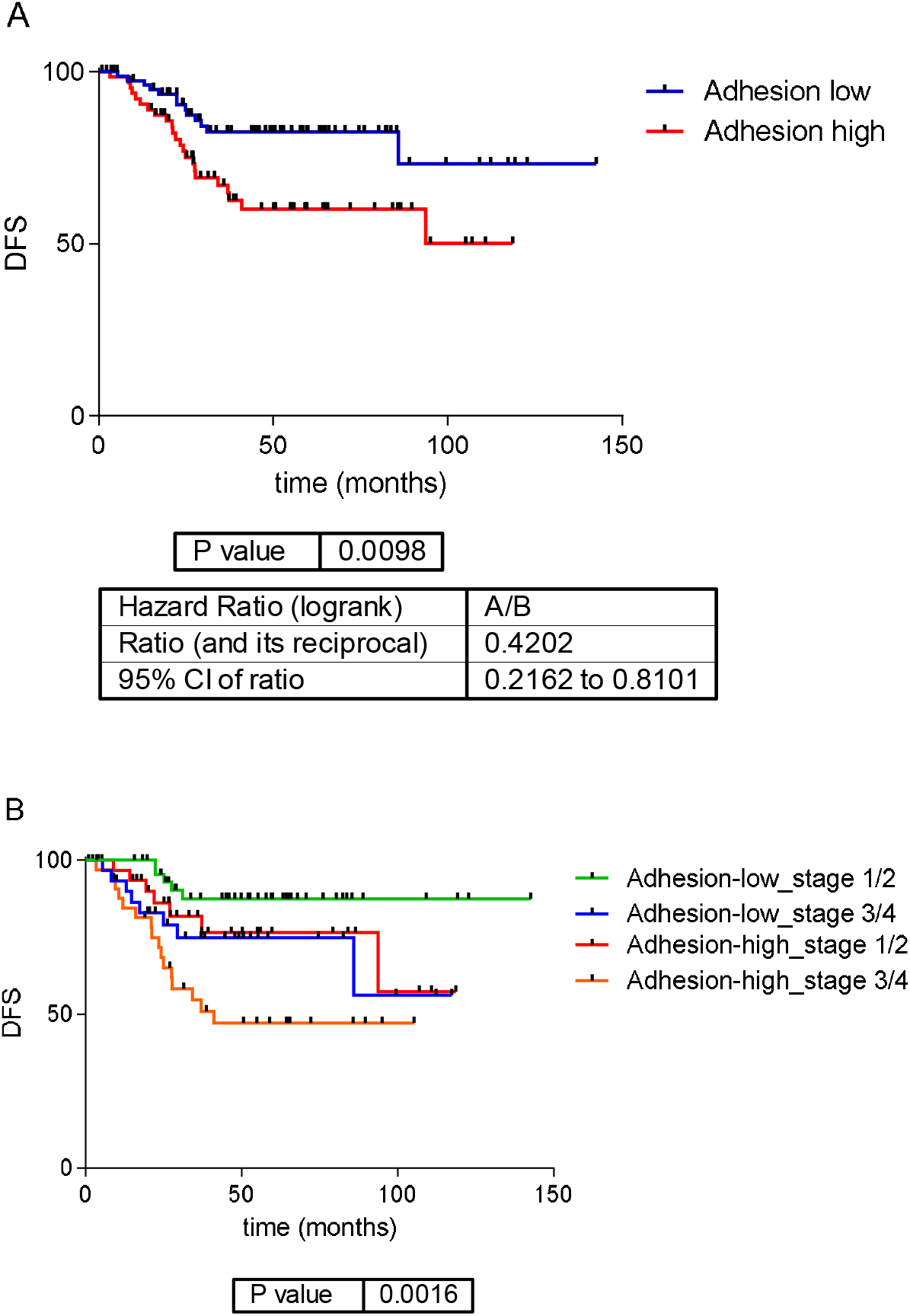
A. Survival analysis of the adhesion groups. Low adhesion tumors showed a significantly better prognosis than high adhesion ones. B. Survival curves of the adhesion groups by tumor stage. DFS: disease free survival.

In order to make a deeper characterization, differences between high and low adhesion groups were evaluated by functional node activity, as defined by the PGM (Sup figure 2). High adhesion group had higher activity of adhesion, immune response, inflammatory response, stem cells and extracellular matrix functional nodes. Meanwhile, low adhesion group presented higher activity of Wnt signaling pathway, plasmatic membrane, transport, metabolism and regulation of cardiac conduction functional nodes.

### 3.6. Immune layer

As for the adhesion layer, the final immune layer was built merging genes from layers third and eighth (immune response and inflammatory response) (Supplementary table 3). The CC determined that the immune layer should be divided into two groups: immune 1, renamed as immune high group, with 364 tumors showing higher expression of genes related to immune response, and immune 2, renamed as immune low group, including 441 tumors with lower expression of these genes. Differences in gene expression between both groups were evaluated using SAM (Figure 4A). Most of the differential genes belonged to the human leukocyte antigen (HLA) complex gene family (Supplementary table 4). The immune layer had no prognostic value in our series (p=0.57, HR=0.82, 95%CI= (0.42-1.59))(Sup figure 3). No differences according to the distribution of tumor location and stage in each immune group were found.

**Figure 4:**
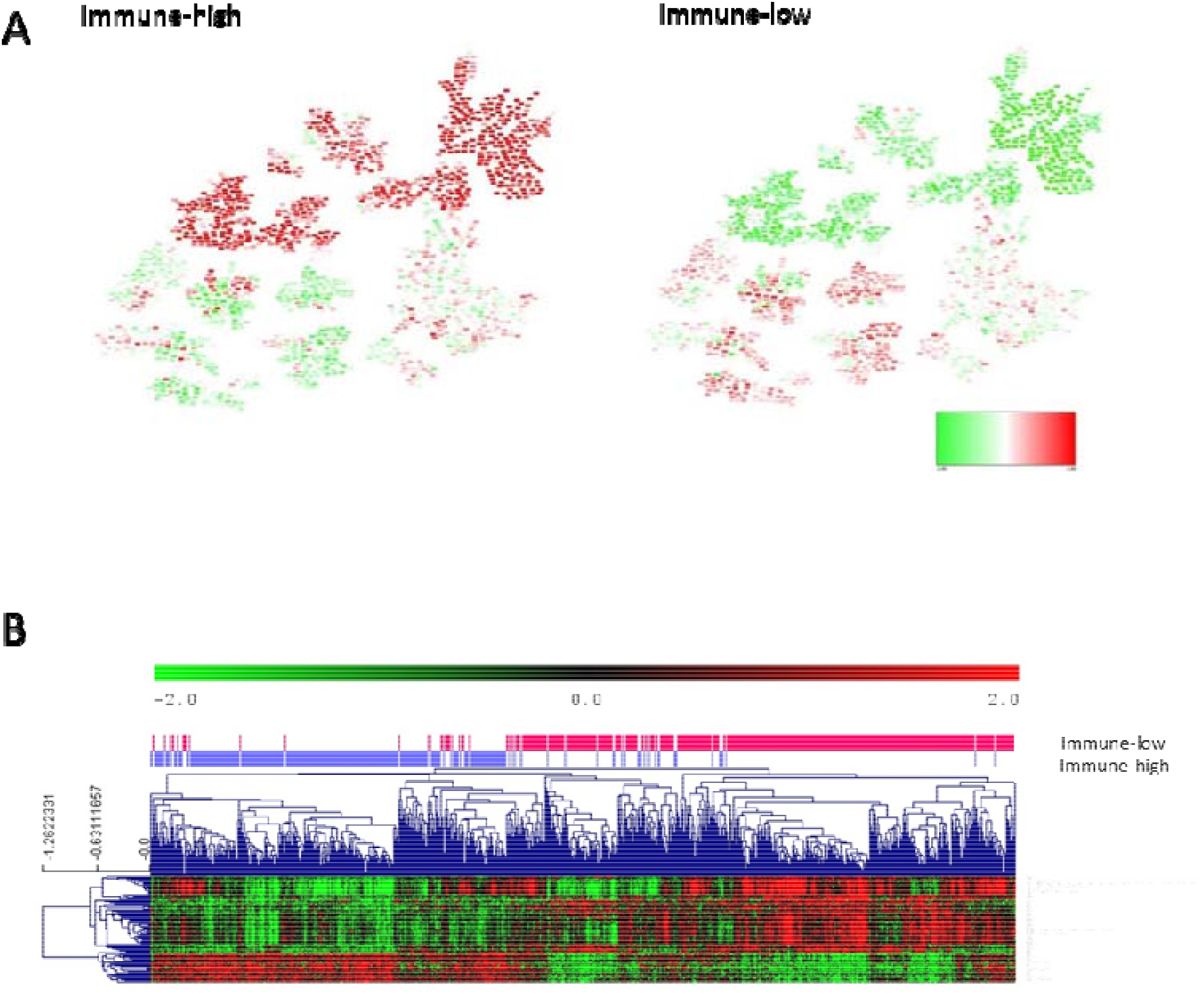
A. Heatmap of the mean gene expression of immune groups in the PGM. Red= overexpressed. Green= underexpressed. B. Differential 100 genes between immune groups identified by SAM.

Functional node activity analysis showed that tumors in the immune high group had a higher expression of genes in the immune, inflammatory, cellular adhesion and extracellular matrix functional nodes. On the contrary, immune low tumors had a higher expression of genes located in the transport, Wnt signaling pathways, nucleus, and regulation of cardiac conduction functional nodes (Figure 4, Sup figure 4 and supplementary table 4).

As viral mimicry response has gained relevance in the last years in cancer related to immune response activation [31], we studied the expression of the genes involved in viral mimicry response in the two immune groups. These immune groups presented a differential expression of the genes involved in viral mimicry response, being higher in the immune-high group (Figure 5).

**Figure 5:**
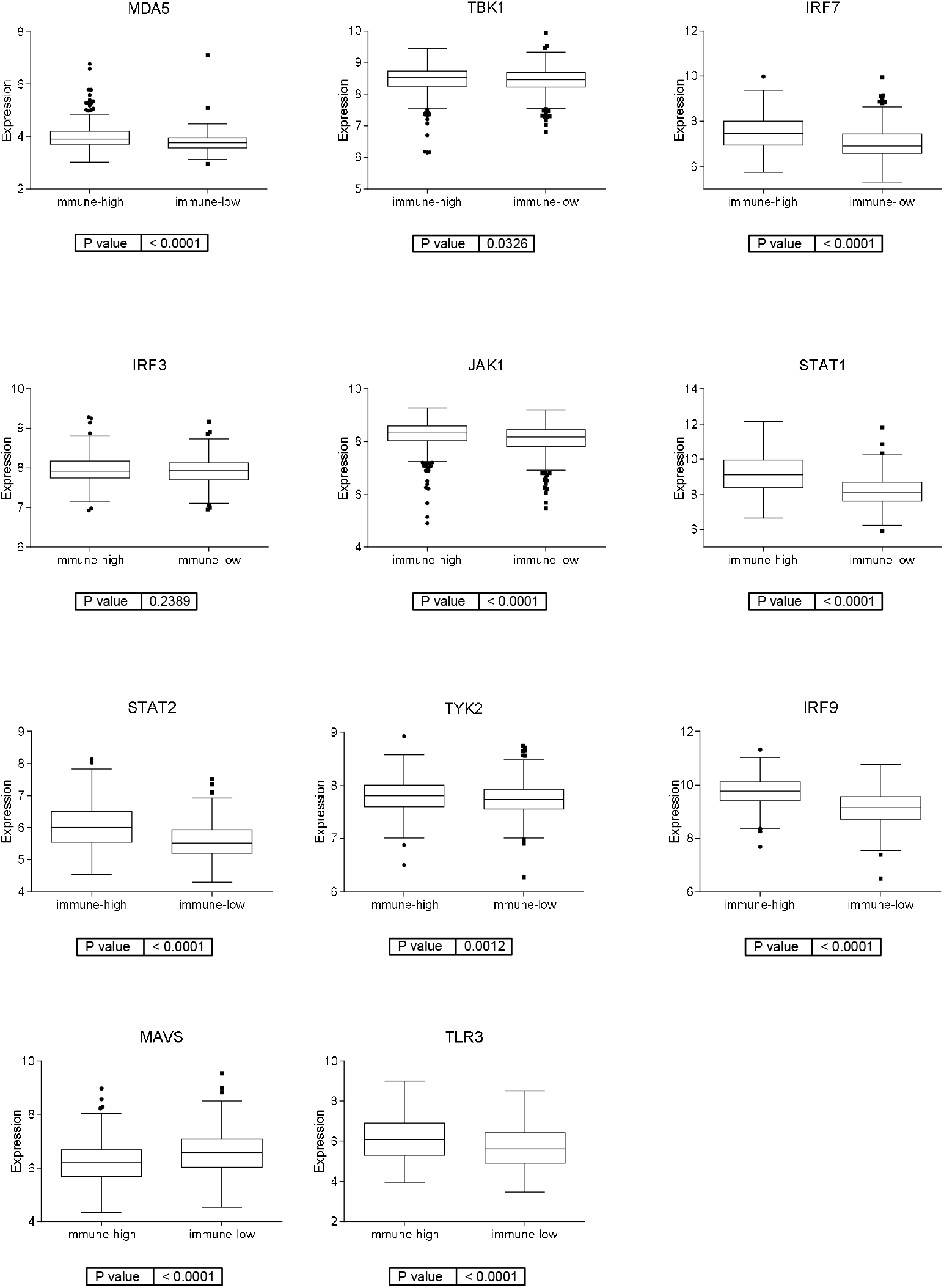
Expression of genes involved in viral mimicry response in the two immune groups.

### 3.7. Molecular layer

The molecular layer resulted from merging the second, fourth and sixth layers and it was divided into four groups by CC. Differences between molecular groups were evaluated using the PGM and the node activities. These analyses showed that molecular group 1 had a higher activity in stem cells, nucleus, regulation of cardiac conduction and transport nodes; molecular group 2 had a higher activity of metabolism node; molecular group 3 had a higher activity of nucleus and Wnt signaling pathways nodes; and molecular group 4 had a higher activity of cell adhesion, extracellular matrix and extracellular response nodes. Therefore, molecular group 1 (221 tumors, 27%) which presented the highest activity for the stem cell functional node, was designed as Stem Cell group. Molecular group 2 (137 tumors, 17%), which had the highest activity of metabolism node, was named as Metabolic group. Molecular group 3 (300 tumors, 37%), which had the highest activity of Wnt signaling pathway node, was named as Wnt pathway group. Finally, molecular group 4 (147 tumors, 18%), which had the highest activity of extracellular response and extracellular matrix nodes, was named Extracellular group (Figure 6A, Sup Fig 5).

**Figure 6:**
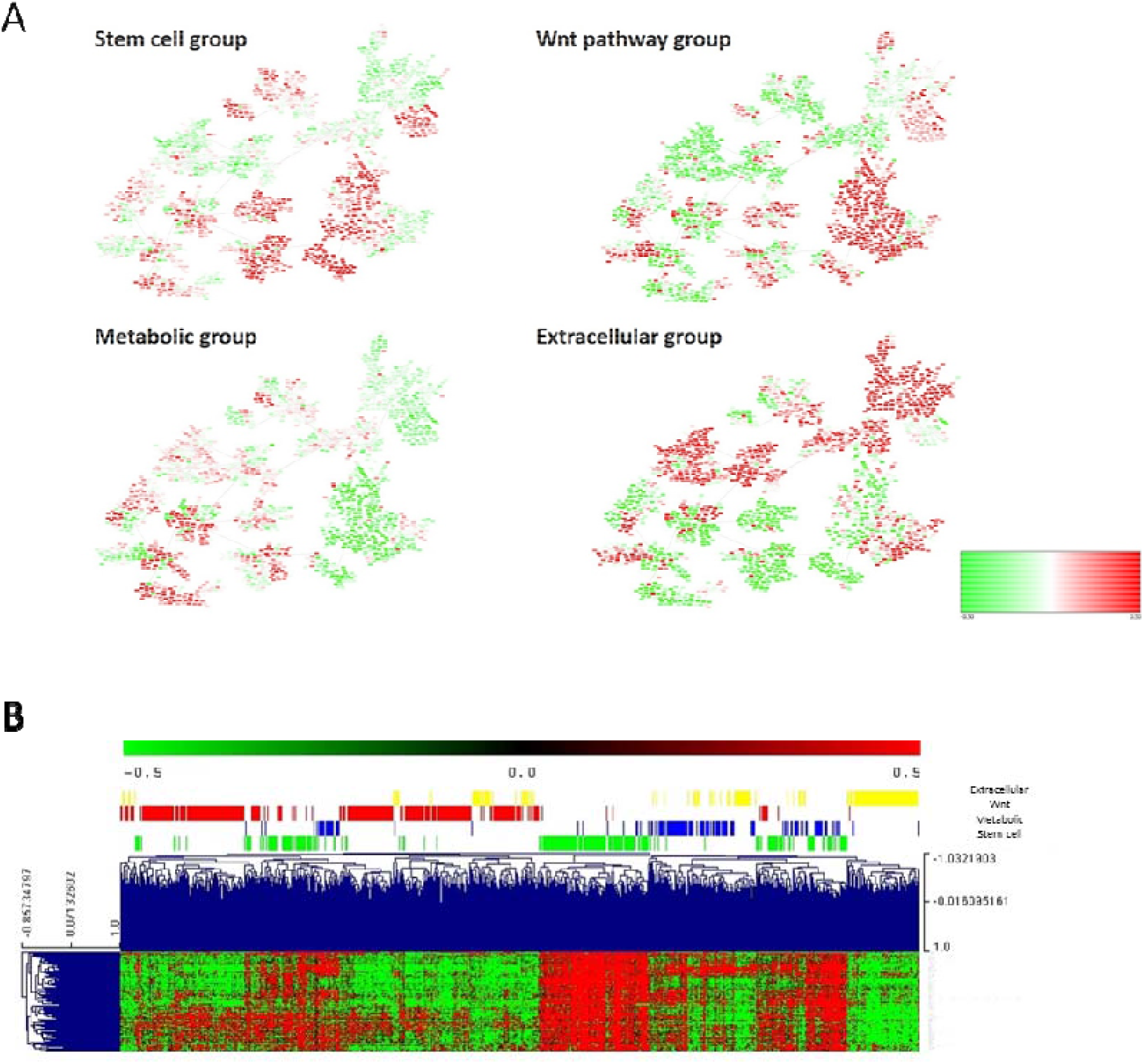
**A**. Heatmap of the mean gene expression of the four molecular groups in the PGM. Red=overexpressed. Green= underexpressed. B. Differential 100 between the four molecular groups.

Stem cell node contained genes involved in stem cell maintenance such as VANGL2 or PBX1. Metabolic node was formed by genes directly involved in metabolism as PHGD or PSAT1 and other genes such as CTSE or REG4. Wnt pathway node contained genes involved in Wnt signaling pathway: RNF43, DKK4, LRP4, AXIN2, etc. Extracellular matrix node was mainly formed by collagens. Regarding the association of the defined molecular groups with clinical parameters, tumor location was distributed significantly different between molecular subtypes (p<0.0001), being Stem cell and Wnt groups mainly composed by distal tumors and metabolic and extracellular groups by proximal tumors. No differences regarding the distribution of tumor stage across molecular groups were found.

Differentially expressed genes between these molecular groups were identified using SAM (figure 6B and supplementary table 4). Stem cell group had also an overexpression of genes related to fatty acid metabolism such as UGT1A1 or UGT1A5, and mucins. Metabolic group had an overexpression of genes involved in metabolic pathways, including cholesterol or tryptophan metabolism. Wnt group had also an overexpression of genes involved in retinol metabolism or epidermal growth factor receptor binding among others. Extracellular group had an overexpression of plasma membrane genes.

The survival analysis of these four groups showed no significant differences, however extracellular group had a better prognosis than the other 3 groups (p=0.0086, HR=0.41 95%CI: (0.13-0.73)) (Sup Figure 6).

### 3.8. Comparison between layer classification and CMS

Once we obtained three independent classifications, we compared them with the CMS. Patients belonging to the CMS1 and CMS4 were mostly immune high whereas CMS2 and CMS3 patients were mostly immune low. The adhesion layer divided the CMS1 patients by half and most of the CMS4 patients were included in the high adhesion group whereas CMS2 and CMS3 were included in the low adhesion group. According to the molecular layer, most of the CMS1 patients belonged to the extracellular molecular subtype, the CMS2 to the Wnt pathway and the CMS3 to the metabolic group (Figure 7, Sup Table 5).

**Figure 7:**
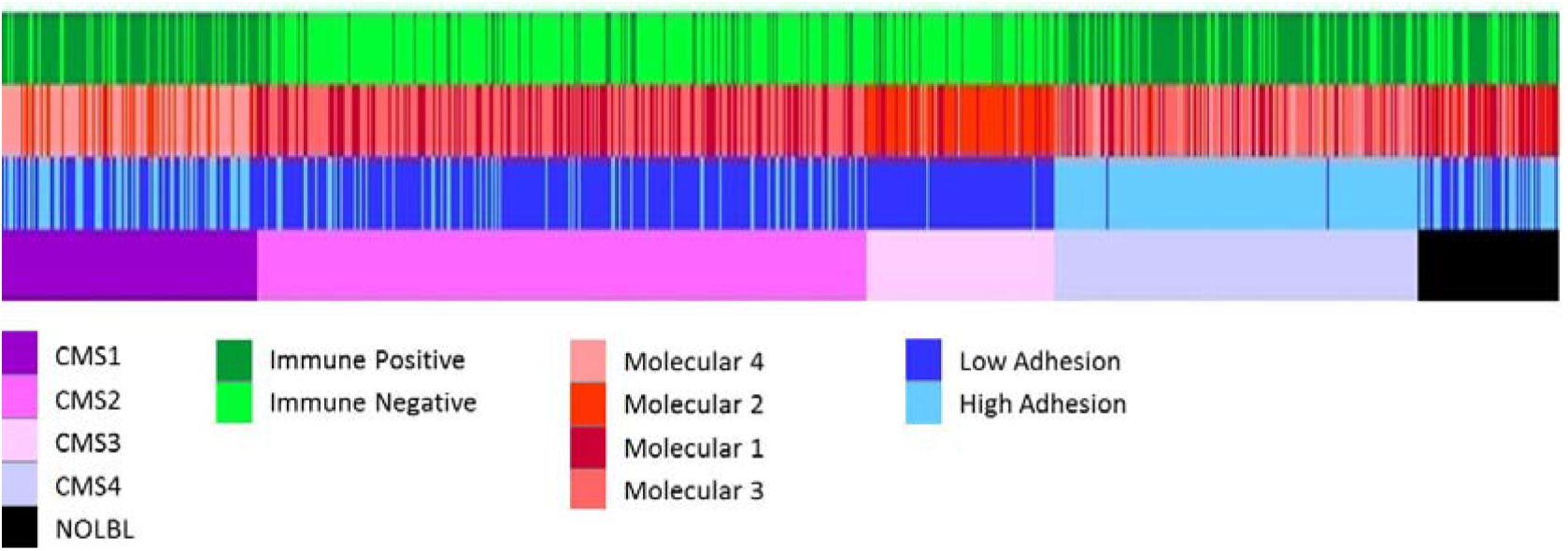
All the classifications of CRC tumors. From top to bottom, immune layer, molecular layer, adhesion layer and CMS classification groups.

## 4. Discussion

Colorectal cancer is a molecularly and clinically heterogeneous disease with high rates of incidence and mortality [1]. Both CRC incidence and mortality are expected to increase in the coming years [32]. Therefore, it is essential to explore new molecular markers and therapeutic applications to improve the prognosis and clinical management of this type of tumor. In order to solve the problems derived from the heterogeneity of colorectal cancer, the international CRCSC (Colorectal Cancer Subtyping Consortium) was created, where colon cancer was classified into four consensus molecular subtypes (CMS) [7]. The prognostic value of CMS classification has been proven in metastatic CRC [33-35] and recent meta-analysis studies find that prognostic and predictive value of the CMS are robust [6], but at the present time, CMS classification has no direct impact on clinical decision-making [12]. Many studies have tried to improve the CMS classification for more refined prognosis predictions [36-39]. However, discovery of new CRC patient stratification methods are still necessary for enhanced diagnosis of CRC, screening for novel therapeutic targets, and improving prognostic tools for CRC.

PGMs have demonstrated their utility in the analysis of tumor omics data, being able to structure molecular information from a functional point of view [13, 14, 40]. Additionally, in other tumor types, such as bladder cancer, Sparse K-means-CC analysis provided independent layers of information from the molecular characteristics of the tumors, for instance, immune information [13, 18, 26]. Therefore, the generation of a classification using this novel approach based on the existence of different informative layers could help to translate into the clinical practice the molecular information generated in the context of CRCSC. This approach allows us the identification of three different levels of information: the adhesion layer, the immune layer and the molecular layer.

The adhesion layer has been divided into two groups, high and low adhesion, and had prognostic value, having the group of patients with low adhesion the best prognosis. Distant CRC metastatic tumor formation is considered to be strongly influenced by the stable adhesion of cancer cells to the small blood vessel walls [41]. In recent years, several studies have shown that adhesion molecules are responsible for tumor progression and metastasis in colorectal cancer, however, the prognostic significance of these markers remains controversial [42]. Also, other adhesion proteins, like FLRT2 and AMIGO2, overexpressed in high adhesion group and located in the adhesion and extracellular matrix nodes respectively, have been suggested to be useful biomarkers for the long-term prognosis of CRC patients [43, 44]. As adhesion genes, extracellular matrix genes were also overexpressed in the adhesion-high group. One of this genes was collagen triple helix repeat containing 1 (CTHRC1), related to an increase in cell migration, motility and invasion. CTHRC1 overexpression was related to poor prognosis of CRC patients and has been defined as a potential diagnostic and prognostic biomarker for patients with CRC [45, 46].

Immunotherapy relies on harnessing the body’s immune system to kill cancer cells [47] and it has revolutionized the treatment of several cancers. Immunotherapy has also shown impressive results in the context of CRC. Patients with mismatch repair (dMMR)/microsatellite-instability-high (MSI-H) metastatic CRC has been observed to have a prolonged benefit to immune checkpoint inhibitors. Consequently, pembrolizumab and nivolumab +/− ipilimumab have obtained the Food and Drug Administration approval for MSI-H/dMMR metastatic CRC [48-50]. The immune layer divided CRC tumors into two groups, immune-high and immune-low. A classification capable of identifying immune-related differences, independent of molecular subtype, may identify tumors that will be good responders to immunotherapy. HLA complex genes were overexpressed in the immune-high group. Tumor cells may escape T cell attack through HLA downregulation [51], so overexpression of HLA complex genes matches with considering this group of patients as optimal candidates for immunotherapy.

Immune-high group had also an overexpression of genes involved in viral mimicry response. Viral mimicry response is a cellular state of active viral response triggered by endogenous stimuli instead of viral infection, in the case of cancer, triggered by retrotransposons [31]. Viral mimicry interprets these retrotransposons as a viral infection and activates interferon response. Viral mimicry response also increases adaptive immune response through the increased expression of antigen processing components and increased expression of retrotransposon-derived peptides [52].

Several of the immune genes in which immune classification was based, were involved in antigen processing and presentation process, and they may be the cause of the activation of viral mimicry response. Moreover, cytidine analogues, azacytidine or decitabine, at low doses, have demonstrated anti-tumor efficacy in colorectal cancer cells by inducing viral mimicry [53] and also it has been demonstrated that they enhance the response to immune checkpoint inhibitors [54]. Therefore, a combination of viral mimicry-related drugs and immunotherapy could be an option for the immune-high patients.

The molecular layer has been divided into 4 groups: stem cells, Wnt pathway, metabolic and extracellular. Cancer stem cells (CSCs) can regulate cancer invasion, distant metastases, and therapy resistance in CRC, as well as contribute to the cancer recurrence of patients [55]. The stem cell subtype presented high expression of genes such as VANGL2 or PBX1, whose function was related to stem cell maintenance. Therefore, these biomarkers could be a possible avenue of study since colorectal cancer stem cells differ from normal stem cells in their tumorigenic potential and susceptibility to chemotherapeutic drugs [56] which would explain the high percentage of relapses in patients with this type of cancer.

Wnt signaling pathway plays an important role in the pathogenesis of CRC [57]. Tumors of the Wnt molecular subtype presented high expression of genes such as RNF43, related to alterations in the Wnt signaling pathway [58]. RNF43 encodes an E3 ubiquitin ligase that negatively regulates Wnt signaling and it is mutated in more than 18% of colorectal adenocarcinomas and endometrial carcinomas. Mutations in RNF43 have clinical relevance because implicates novel therapeutic options in CRC. Preclinical studies have shown that mutations in RNF43 make Wnt-induced cancer cells susceptible to pharmacological inhibition of Wnt signaling by porcupine. Porcupine is an O-acetyltransferase that is part of the Wnt pathway, and could be postulated as a possible therapy in this type of Wnt-induced tumors [59-61]. To date, five porcupine inhibitors have entered phase I/II clinical trials in patients with advanced solid tumors and showed promising preliminary clinical data [62, 63]. Porcupine inhibitors were well-tolerated, also when they were used in combination with anti-PD-1 therapy [64]

In tumors of the metabolic molecular group, a high expression of genes that play an essential role in cancer-specific metabolic reprogramming, such as PHGDH, has been observed [65]. PHGDH is a metabolic enzyme involved in the serine synthetic pathway and it appears to play a central role in supporting cancer growth and proliferation so it is a promising drug target for cancer therapy. Different PHGDH inhibitors have been reported but currently they have not yet led to the development of compounds that can be therapeutically used [65]. Other gene overexpressed in the metabolic group was phosphoserine aminotransferase 1 (PSAT1), a gene related to serine biosynthesis. Certain studies concluded that overexpression of PSAT1 is significantly associated with resistance to chemotherapy with irinotecan, 5-fluorouracil and leucovorin, so the inhibition of this gene could prevent patients of this group from developing resistance to chemotherapy [66]. Other genes involved in metabolism such as REG4 and CTSE, were also overexpressed in the metabolic subtype and they have been previously related to CRC prognosis. Regenerating islet-derived type 4 (REG4) is a member of the calcium-dependent lectin gene superfamily and it was associated with a relatively unfavourable prognosis in various cancers including CRC [67, 68]. Cathepsin E (CTSE) is an adverse prognostic factor for survival among rectal cancer patients receiving chemo-radiotherapy [69]. Cathepsins have been implicated to play a role in the invasion and metastasis of colorectal cancer. Inhibitors targeting some cathepsins like S and K are already in clinical evaluation [70] and inhibition of the Reg4-CD44/CD44ICD pathway has been proposed as a future therapeutic target for colon cancer patients [71]. The use of REG4 and CTSE inhibitors could be a targeted treatment for patients of this molecular group.

The extracellular molecular group was characterized by a high expression of collagens, which could be one of the reasons why it is the subtype with the worst prognosis. Among extracellular matrix, adhesive components type I collagen is one of the important factors regulating cancer-related events at different tumorigenesis stages [72] The COL1A1 gene encodes a pro-α1 chain of type I collagen and it has been demonstrated that is overexpressed in colon cancer and it may be a driving gene for colon cancer progression [73, 74]. Different inhibitors and drugs that regulate collagen biosynthesized processes and collagen distribution arrangement have been described and preclinical studies on collagen-related therapy have demonstrated encouraging outcomes [75]. Patients of this molecular subtype could be candidates for collagen inhibitor therapies.

We compared our three classifications of CRC tumors with the classification through CMS groups. The CMS classification mixed immunological, histological and molecular information. The present study has been able to corroborate some of the molecular characteristics defined in the CMS but it has also been possible to identify two layers of information that were independent for the molecular features of the CRC tumors related to adhesion and the immune status. These different levels of information allowed complementing the molecular characteristics exposed in the CMS and it has also been possible to add new information that allowed patients with different CMS to benefit from the same therapeutic strategy.

For instance, although the CRCSC determined that all CMS1 patients were immune positive [7], our results suggested that 80% of CMS1 patients were immune-high, while 20% of patients in the CMS1 group had low expression of immune response-related genes. This would mean that these patients are not optimal candidates for immunotherapy. Moreover, most CMS2 as CMS3 tumors share the characteristic of having a low expression of immune genes (82% and 81% of patients, respectively), being considered cold tumors that do not respond to immunotherapy [76]. However, our molecular classification showed that 18% of CMS2 patients and 19% of CMS3 patients had a high immune status so they could be candidates for immunotherapy. Therefore, an analysis based on different biological layers allow a more accurate classification of CRC patients according to their immune status independently of the CMS group to which they belong. The molecular characterization obtained using the described analysis tools provided complementary information to that of the CMS group classification that may have important implications for the choice of treatment for each patient, such as immunotherapy.

As for the CMS2 group, CRCSC exposed that this group presents a close similarity to the classical model of CRC carcinogenesis with activation of the WNT and MYC signalling pathways [7]. Although our molecular classification divided CMS2 patients into the Wnt molecular subtype and the stem cell molecular subtype, both subtypes presented high functional activity of the Wnt signalling pathway-related node and both could benefit from porcupine Wnt pathway inhibition therapy.

As for the CMS3 group, the CRCSC established that only patients in this group could benefit from possible therapies with PHGHD and PSAT1 or other metabolism-related molecules. 24.7% of CMS3 patients do not correspond to the metabolic molecular subtype, so other therapeutic options should be explored for these patients since, as seen in this study, they are not characterized by a high expression of genes related to metabolism even though they have been included in CMS3.

Regarding the CMS4 group, our analysis determined that 74% of CMS4 patients were immune high. This fact is consistent with the CRCSC classification that described the relationship between CMS4 patients with the presence of high infiltration of cytotoxic T cells [77] and high expression of immune genes [7, 37] so, as with CMS1, these patients could be candidates for immunotherapy [78]. However, in this study, it has been possible to determine that 26% of CMS4 patients are immune low and therefore would not be good candidates for immunotherapy.

On the other hand, CMS4 is the subtype with the worst prognosis [7]. Regarding the adhesion subtype, 98% of CMS4 patients belong to the high adhesion group and this is consistent with the survival analysis that determined that the adhesion layer had prognostic value, showing worse prognostic in high adhesion tumors.

To sum up, this study allows minimizing the percentage of patients without a specific treatment since the layer classification allows adding information about immune and adhesion status. Thus, patients of the stem cell molecular subtype and the Wnt molecular subtype could benefit from porcupine inhibition therapy, patients of the metabolic molecular subtype from possible therapies related with REG4 and CTSE, and patients of the extracellular molecular subtype from possible therapies related to COL1A1. On the other hand, the classification of patients by immune subtype, independent of CMS, provides valuable information to select the most suitable patients for immunotherapy treatment and viral mimicry therapies.

The study has some limitations. First, a validation of all the obtained classifications in an independent CRC cohort is needed. In addition, a validation of the proposed therapeutic strategies for each group in cell cultures or murine models should be performed. Moreover, only 12% of the tumors were stage IV, so there were underrepresented, as happened in the CMS study. However, this is the group where possible molecular targets are most interesting because of their potential therapeutic utility. The study is a retrospective cohort, prior to immunotherapy administration which may change the prognosis of some patients, especially those with microsatellite instability. Finally, these groups should be study in the context of other clinical biomarkers such as RAS/RAF or microsatellite instability.

## Conclusions

In conclusion, the generation of a classification of colorectal cancer according to the different biological realities of the tumor using probabilistic graphic models and layer analysis allows the identification of four molecular subtypes of colorectal cancer and established two extra independent classification based on adhesion and immune features, respectively. These classifications may help researchers and clinicians to search for new therapeutic targets and more specific treatments.

## Data Availability

All data produced in the present work are contained in the manuscript.

## FUNDING

EL-C is supported by the Spanish Economy and Competitiveness Ministry (PTQ2018-009760). This research was funded by Jesús Antolín Garciarena fellowship from IdiPAZ

## AUTHOR CONTRIBUTIONS

DM-P, AG-P, JAFV, JF, and LTF: Conceptualization. EL-C, GP-V, MF-G, SL-A, RL-V, MD-A, AG-P, DM-P, JAFV, JF and LT-F: Formal analyses. LT-F: Supervision. EL-C: Writing original draft. LT-F: Writing-review and editing.

## CONFLICT OF INTERESTS

AG-P and JAFV are shareholders of Biomedica Molecular Medicine SL. EL-C is an employee of Biomedica Molecular Medicine. The other authors declare that there are no conflicts of interest.

## Notes

### Funding Statement

EL-C is supported by the Spanish Economy and Competitiveness Ministry (PTQ2018-009760). This research was funded by Jesus Antolin Garciarena fellowship from IdiPAZ

### Author Declarations

Three colorectal tumor gene expression databases (GSE17536, GSE35896, and GSE39582) were analyzed. Data is available at synapse platform (https://www.synapse.org/#!Synapse:syn2623706/wiki/67246).

## References

1. Sung H, Ferlay J, Siegel RL, Laversanne M, Soerjomataram I, Jemal A, et al. Global Cancer Statistics 2020: GLOBOCAN Estimates of Incidence and Mortality Worldwide for 36 Cancers in 185 Countries. CA Cancer J Clin. 2021;71(3):209–49. Epub 20210204. doi: 10.3322/caac.21660. PubMed PMID: 33538338.

2. Comprehensive molecular characterization of human colon and rectal cancer. Nature. 2012;487(7407):330–7. Epub 20120718. doi: 10.1038/nature11252. PubMed PMID: 22810696; PubMed Central PMCID: PMCPMC3401966.

3. Lee GH, Malietzis G, Askari A, Bernardo D, Al-Hassi HO, Clark SK. Is right-sided colon cancer different to left-sided colorectal cancer? - a systematic review. Eur J Surg Oncol. 2015;41(3):300–8. Epub 20141113. doi: 10.1016/j.ejso.2014.11.001. PubMed PMID: 25468456.

4. Bufill JA. Colorectal cancer: evidence for distinct genetic categories based on proximal or distal tumor location. Ann Intern Med. 1990;113(10):779–88. doi: 10.7326/0003-4819-113-10-779. PubMed PMID: 2240880.

5. Gong P, Chen C, Wang Z, Zhang X, Hu W, Hu Z, et al. Prognostic significance for colorectal carcinoid tumors based on the 8th edition TNM staging system. Cancer Med. 2020;9(21):7979–87. Epub 20200908. doi: 10.1002/cam4.3431. PubMed PMID: 32897004; PubMed Central PMCID: PMCPMC7643648.

6. Ten Hoorn S, de Back TR, Sommeijer DW, Vermeulen L. Clinical Value of Consensus Molecular Subtypes in Colorectal Cancer: A Systematic Review and Meta-Analysis. J Natl Cancer Inst. 2022;114(4):503–16. doi: 10.1093/jnci/djab106. PubMed PMID: 34077519; PubMed Central PMCID: PMCPMC9002278.

7. Guinney J, Dienstmann R, Wang X, de Reyniès A, Schlicker A, Soneson C, et al. The consensus molecular subtypes of colorectal cancer. Nat Med. 2015;21(11):1350–6. Epub 20151012. doi: 10.1038/nm.3967. PubMed PMID: 26457759; PubMed Central PMCID: PMCPMC4636487.

8. Mooi JK, Wirapati P, Asher R, Lee CK, Savas P, Price TJ, et al. The prognostic impact of consensus molecular subtypes (CMS) and its predictive effects for bevacizumab benefit in metastatic colorectal cancer: molecular analysis of the AGITG MAX clinical trial. Ann Oncol. 2018;29(11):2240–6. doi: 10.1093/annonc/mdy410. PubMed PMID: 30247524.

9. Lenz HJ, Ou FS, Venook AP, Hochster HS, Niedzwiecki D, Goldberg RM, et al. Impact of Consensus Molecular Subtype on Survival in Patients With Metastatic Colorectal Cancer: Results From CALGB/SWOG 80405 (Alliance). J Clin Oncol. 2019;37(22):1876–85. Epub 20190501. doi: 10.1200/jco.18.02258. PubMed PMID: 31042420; PubMed Central PMCID: PMCPMC6675593.

10. Okita A, Takahashi S, Ouchi K, Inoue M, Watanabe M, Endo M, et al. Consensus molecular subtypes classification of colorectal cancer as a predictive factor for chemotherapeutic efficacy against metastatic colorectal cancer. Oncotarget. 2018;9(27):18698–711. Epub 20180410. doi: 10.18632/oncotarget.24617. PubMed PMID: 29721154; PubMed Central PMCID: PMCPMC5922348.

11. Rodriguez-Salas N, Dominguez G, Barderas R, Mendiola M, García-Albéniz X, Maurel J, et al. Clinical relevance of colorectal cancer molecular subtypes. Crit Rev Oncol Hematol. 2017;109:9–19. Epub 20161123. doi: 10.1016/j.critrevonc.2016.11.007. PubMed PMID: 28010901.

12. Ignatova EO, Kozlov E, Ivanov M, Mileyko V, Menshikova S, Sun H, et al. Clinical significance of molecular subtypes of gastrointestinal tract adenocarcinoma. World J Gastrointest Oncol. 2022;14(3):628–45. doi: 10.4251/wjgo.v14.i3.628. PubMed PMID: 35321271; PubMed Central PMCID: PMCPMC8919013.

13. de Velasco G, Trilla-Fuertes L, Gamez-Pozo A, Urbanowicz M, Ruiz-Ares G, Sepúlveda JM, et al. Urothelial cancer proteomics provides both prognostic and functional information. Sci Rep. 2017;7(1):15819. Epub 20171117. doi: 10.1038/s41598-017-15920-6. PubMed PMID: 29150671; PubMed Central PMCID: PMCPMC5694001.

14. Gámez-Pozo A, Berges-Soria J, Arevalillo JM, Nanni P, López-Vacas R, Navarro H, et al. Combined Label-Free Quantitative Proteomics and microRNA Expression Analysis of Breast Cancer Unravel Molecular Differences with Clinical Implications. Cancer Res. 2015;75(11):2243–53. Epub 2015/04/18. doi: 10.1158/0008-5472.can-14-1937. PubMed PMID: 25883093.

15. Gámez-Pozo A, Trilla-Fuertes L, Berges-Soria J, Selevsek N, López-Vacas R, Díaz-Almirón M, et al. Functional proteomics outlines the complexity of breast cancer molecular subtypes. Sci Rep. 2017;7(1):10100. Epub 2017/09/01. doi: 10.1038/s41598-017-10493-w. PubMed PMID: 28855612; PubMed Central PMCID: PMCPMC5577137 L.T.-F. is an employee of Biomedica Molecular Medicine S.L. The other authors declare that they have no competing interests.

16. Trilla-Fuertes L, Gámez-Pozo A, Arevalillo JM, Díaz-Almirón M, Prado-Vázquez G, Zapater-Moros A, et al. Molecular characterization of breast cancer cell response to metabolic drugs. Oncotarget. 2018;9(11):9645–60. Epub 2018/03/09. doi: 10.18632/oncotarget.24047. PubMed PMID: 29515760; PubMed Central PMCID: PMCPMC5839391.

17. Zapater-Moros A, Gámez-Pozo A, Prado-Vázquez G, Trilla-Fuertes L, Arevalillo JM, Díaz-Almirón M, et al. Probabilistic graphical models relate immune status with response to neoadjuvant chemotherapy in breast cancer. Oncotarget. 2018;9(45):27586–94. Epub 20180612. doi: 10.18632/oncotarget.25496. PubMed PMID: 29963222; PubMed Central PMCID: PMCPMC6021258.

18. Prado-Vázquez G, Gámez-Pozo A, Trilla-Fuertes L, Arevalillo JM, Zapater-Moros A, Ferrer-Gómez M, et al. A novel approach to triple-negative breast cancer molecular classification reveals a luminal immune-positive subgroup with good prognoses. Sci Rep. 2019;9(1):1538. Epub 20190207. doi: 10.1038/s41598-018-38364-y. PubMed PMID: 30733547; PubMed Central PMCID: PMCPMC6367406.

19. Witten DM, Tibshirani R. A framework for feature selection in clustering. J Am Stat Assoc. 2010;105(490):713–26. doi: 10.1198/jasa.2010.tm09415. PubMed PMID: 20811510; PubMed Central PMCID: PMCPMC2930825.

20. Monti S, Tamayo P, Mesirov J, Golub T. Consensus Clustering: A Resampling-Based Method for Class Discovery and Visualization of Gene Expression Microarray Data. Machine Learning. 2003;52(1):91–118. doi: 10.1023/A:1023949509487.

21. Ritchie ME, Phipson B, Wu D, Hu Y, Law CW, Shi W, et al. limma powers differential expression analyses for RNA-sequencing and microarray studies. Nucleic Acids Res. 2015;43(7):e47. Epub 20150120. doi: 10.1093/nar/gkv007. PubMed PMID: 25605792; PubMed Central PMCID: PMCPMC4402510.

22. Derry JM, Mangravite LM, Suver C, Furia MD, Henderson D, Schildwachter X, et al. Developing predictive molecular maps of human disease through community-based modeling. Nat Genet. 2012;44(2):127–30. Epub 20120127. doi: 10.1038/ng.1089. PubMed PMID: 22281773; PubMed Central PMCID: PMCPMC3643818.

23. Abreu GCG, Labouriau R, Edwards D. High-Dimensional Graphical Model Search with the gRapHD R Package. Journal of Statistical Software. 2010;37(1):1 – 18. doi: 10.18637/jss.v037.i01.

24. Computing R. R: A language and environment for statistical computing. Vienna: R Core Team. 2013.

25. Huang da W, Sherman BT, Lempicki RA. Systematic and integrative analysis of large gene lists using DAVID bioinformatics resources. Nat Protoc. 2009;4(1):44–57. doi: 10.1038/nprot.2008.211. PubMed PMID: 19131956.

26. Trilla-Fuertes L, Gámez-Pozo A, Prado-Vázquez G, Zapater-Moros A, Díaz-Almirón M, Arevalillo JM, et al. Biological molecular layer classification of muscle-invasive bladder cancer opens new treatment opportunities. BMC Cancer. 2019;19(1):636. Epub 2019/06/30. doi: 10.1186/s12885-019-5858-z. PubMed PMID: 31253132; PubMed Central PMCID: PMCPMC6599340.

27. Tusher VG, Tibshirani R, Chu G. Significance analysis of microarrays applied to the ionizing radiation response. Proc Natl Acad Sci U S A. 2001;98(9):5116–21. Epub 20010417. doi: 10.1073/pnas.091062498. PubMed PMID: 11309499; PubMed Central PMCID: PMCPMC33173.

28. Saeed AI, Sharov V, White J, Li J, Liang W, Bhagabati N, et al. TM4: a free, open-source system for microarray data management and analysis. Biotechniques. 2003;34(2):374–8. doi: 10.2144/03342mt01. PubMed PMID: 12613259.

29. Shannon P, Markiel A, Ozier O, Baliga NS, Wang JT, Ramage D, et al. Cytoscape: a software environment for integrated models of biomolecular interaction networks. Genome Res. 2003;13(11):2498–504. doi: 10.1101/gr.1239303. PubMed PMID: 14597658; PubMed Central PMCID: PMCPMC403769.

30. Edgar R, Domrachev M, Lash AE. Gene Expression Omnibus: NCBI gene expression and hybridization array data repository. Nucleic Acids Res. 2002;30(1):207–10. doi: 10.1093/nar/30.1.207. PubMed PMID: 11752295; PubMed Central PMCID: PMCPMC99122.

31. Chen R, Ishak CA, De Carvalho DD. Endogenous Retroelements and the Viral Mimicry Response in Cancer Therapy and Cellular Homeostasis. Cancer Discov. 2021;11(11):2707–25. Epub 20211014. doi: 10.1158/2159-8290.CD-21-0506. PubMed PMID: 34649957.

32. Xi Y, Xu P. Global colorectal cancer burden in 2020 and projections to 2040. Transl Oncol. 2021;14(10):101174. Epub 20210706. doi: 10.1016/j.tranon.2021.101174. PubMed PMID: 34243011; PubMed Central PMCID: PMCPMC8273208.

33. Rebersek M. Consensus molecular subtypes (CMS) in metastatic colorectal cancer - personalized medicine decision. Radiol Oncol. 2020;54(3):272–7. Epub 20200528. doi: 10.2478/raon-2020-0031. PubMed PMID: 32463385; PubMed Central PMCID: PMCPMC7409603.

34. Lenz HJ, Argiles G, Yoshino T, Tejpar S, Ciardiello F, Braunger J, et al. Association of Consensus Molecular Subtypes and Molecular Markers With Clinical Outcomes in Patients With Metastatic Colorectal Cancer: Biomarker Analyses From LUME-Colon 1. Clin Colorectal Cancer. 2021;20(1):84-95.e8. Epub 20200915. doi: 10.1016/j.clcc.2020.09.003. PubMed PMID: 33041226.

35. Stintzing S, Wirapati P, Lenz HJ, Neureiter D, Fischer von Weikersthal L, Decker T, et al. Consensus molecular subgroups (CMS) of colorectal cancer (CRC) and first-line efficacy of FOLFIRI plus cetuximab or bevacizumab in the FIRE3 (AIO KRK-0306) trial. Ann Oncol. 2019;30(11):1796–803. doi: 10.1093/annonc/mdz387. PubMed PMID: 31868905; PubMed Central PMCID: PMCPMC6927316.

36. Bramsen JB, Rasmussen MH, Ongen H, Mattesen TB, Ørntoft MW, Árnadóttir SS, et al. Molecular-Subtype-Specific Biomarkers Improve Prediction of Prognosis in Colorectal Cancer. Cell Rep. 2017;19(6):1268–80. doi: 10.1016/j.celrep.2017.04.045. PubMed PMID: 28494874.

37. Dienstmann R, Vermeulen L, Guinney J, Kopetz S, Tejpar S, Tabernero J. Consensus molecular subtypes and the evolution of precision medicine in colorectal cancer. Nat Rev Cancer. 2017;17(2):79–92. Epub 20170104. doi: 10.1038/nrc.2016.126. PubMed PMID: 28050011.

38. Adam RS, Poel D, Ferreira Moreno L, Spronck J, de Back TR, Torang A, et al. Development of a miRNA-based classifier for detection of colorectal cancer molecular subtypes. Mol Oncol. 2022. Epub 20220317. doi: 10.1002/1878-0261.13210. PubMed PMID: 35298091.

39. Hu F, Wang J, Zhang M, Wang S, Zhao L, Yang H, et al. Comprehensive Analysis of Subtype-Specific Molecular Characteristics of Colon Cancer: Specific Genes, Driver Genes, Signaling Pathways, and Immunotherapy Responses. Front Cell Dev Biol. 2021;9:758776. Epub 20211129. doi: 10.3389/fcell.2021.758776. PubMed PMID: 34912802; PubMed Central PMCID: PMCPMC8667669.

40. Gámez-Pozo A, Trilla-Fuertes L, Prado-Vázquez G, Chiva C, López-Vacas R, Nanni P, et al. Prediction of adjuvant chemotherapy response in triple negative breast cancer with discovery and targeted proteomics. PLoS One. 2017;12(6):e0178296. Epub 20170608. doi: 10.1371/journal.pone.0178296. PubMed PMID: 28594844; PubMed Central PMCID: PMCPMC5464546.

41. Korb T, Schlüter K, Enns A, Spiegel HU, Senninger N, Nicolson GL, et al. Integrity of actin fibers and microtubules influences metastatic tumor cell adhesion. Exp Cell Res. 2004;299(1):236–47. doi: 10.1016/j.yexcr.2004.06.001. PubMed PMID: 15302590.

42. Seo KJ, Kim M, Kim J. Prognostic implications of adhesion molecule expression in colorectal cancer. Int J Clin Exp Pathol. 2015;8(4):4148–57. Epub 20150401. PubMed PMID: 26097606; PubMed Central PMCID: PMCPMC4466993.

43. Ando T, Tai-Nagara I, Sugiura Y, Kusumoto D, Okabayashi K, Kido Y, et al. Tumor-specific interendothelial adhesion mediated by FLRT2 facilitates cancer aggressiveness. J Clin Invest. 2022;132(6). doi: 10.1172/jci153626. PubMed PMID: 35104247; PubMed Central PMCID: PMCPMC8920344.

44. Goto K, Osaki M, Izutsu R, Tanaka H, Sasaki R, Tanio A, et al. Establishment of an antibody specific for AMIGO2 improves immunohistochemical evaluation of liver metastases and clinical outcomes in patients with colorectal cancer. Diagn Pathol. 2022;17(1):16. Epub 20220130. doi: 10.1186/s13000-021-01176-2. PubMed PMID: 35094710; PubMed Central PMCID: PMCPMC8802484.

45. Meng C, Zhang Y, Jiang D, Wang J. CTHRC1 is a prognosis-related biomarker correlated with immune infiltrates in colon adenocarcinoma. World J Surg Oncol. 2022;20(1):89. Epub 20220320. doi: 10.1186/s12957-022-02557-7. PubMed PMID: 35307012; PubMed Central PMCID: PMCPMC8934523.

46. Pang C, Wang H, Shen C, Liang H. Application Potential of CTHRC1 as a Diagnostic and Prognostic Indicator for Colon Adenocarcinoma. Front Mol Biosci. 2022;9:849771. Epub 20220301. doi: 10.3389/fmolb.2022.849771. PubMed PMID: 35300110; PubMed Central PMCID: PMCPMC8921526.

47. Papaioannou NE, Beniata OV, Vitsos P, Tsitsilonis O, Samara P. Harnessing the immune system to improve cancer therapy. Ann Transl Med. 2016;4(14):261. doi: 10.21037/atm.2016.04.01. PubMed PMID: 27563648; PubMed Central PMCID: PMCPMC4971375.

48. Gorzo A, Galos D, Volovat SR, Lungulescu CV, Burz C, Sur D. Landscape of Immunotherapy Options for Colorectal Cancer: Current Knowledge and Future Perspectives beyond Immune Checkpoint Blockade. Life (Basel). 2022;12(2). Epub 20220202. doi: 10.3390/life12020229. PubMed PMID: 35207516; PubMed Central PMCID: PMCPMC8878674.

49. Morse MA, Hochster H, Benson A. Perspectives on Treatment of Metastatic Colorectal Cancer with Immune Checkpoint Inhibitor Therapy. Oncologist. 2020;25(1):33–45. Epub 20190805. doi: 10.1634/theoncologist.2019-0176. PubMed PMID: 31383813; PubMed Central PMCID: PMCPMC6964145.

50. Huyghe N, Baldin P, Van den Eynde M. Immunotherapy with immune checkpoint inhibitors in colorectal cancer: what is the future beyond deficient mismatch-repair tumours? Gastroenterol Rep (Oxf). 2020;8(1):11–24. Epub 20191125. doi: 10.1093/gastro/goz061. PubMed PMID: 32104582; PubMed Central PMCID: PMCPMC7034232.

51. Kawazu M, Ueno T, Saeki K, Sax N, Togashi Y, Kanaseki T, et al. HLA Class I Analysis Provides Insight Into the Genetic and Epigenetic Background of Immune Evasion in Colorectal Cancer With High Microsatellite Instability. Gastroenterology. 2022;162(3):799–812. Epub 20211021. doi: 10.1053/j.gastro.2021.10.010. PubMed PMID: 34687740.

52. Jones PA, Ohtani H, Chakravarthy A, De Carvalho DD. Epigenetic therapy in immune-oncology. Nat Rev Cancer. 2019;19(3):151–61. doi: 10.1038/s41568-019-0109-9. PubMed PMID: 30723290.

53. Roulois D, Loo Yau H, Singhania R, Wang Y, Danesh A, Shen SY, et al. DNA-Demethylating Agents Target Colorectal Cancer Cells by Inducing Viral Mimicry by Endogenous Transcripts. Cell. 2015;162(5):961–73. doi: 10.1016/j.cell.2015.07.056. PubMed PMID: 26317465; PubMed Central PMCID: PMCPMC4843502.

54. Loo Yau H, Ettayebi I, De Carvalho DD. The Cancer Epigenome: Exploiting Its Vulnerabilities for Immunotherapy. Trends Cell Biol. 2019;29(1):31–43. Epub 20180825. doi: 10.1016/j.tcb.2018.07.006. PubMed PMID: 30153961.

55. Dalerba P, Clarke MF. Cancer stem cells and tumor metastasis: first steps into uncharted territory. Cell Stem Cell. 2007;1(3):241–2. doi: 10.1016/j.stem.2007.08.012. PubMed PMID: 18371356.

56. Abetov D, Mustapova Z, Saliev T, Bulanin D. Biomarkers and signaling pathways of colorectal cancer stem cells. Tumour Biol. 2015;36(3):1339–53. Epub 20150214. doi: 10.1007/s13277-015-3198-4. PubMed PMID: 25680406.

57. Zhao H, Ming T, Tang S, Ren S, Yang H, Liu M, et al. Wnt signaling in colorectal cancer: pathogenic role and therapeutic target. Mol Cancer. 2022;21(1):144. Epub 20220714. doi: 10.1186/s12943-022-01616-7. PubMed PMID: 35836256; PubMed Central PMCID: PMCPMC9281132.

58. Giannakis M, Hodis E, Jasmine Mu X, Yamauchi M, Rosenbluh J, Cibulskis K, et al. RNF43 is frequently mutated in colorectal and endometrial cancers. Nat Genet. 2014;46(12):1264–6. Epub 20141026. doi: 10.1038/ng.3127. PubMed PMID: 25344691; PubMed Central PMCID: PMCPMC4283570.

59. Ho SY, Keller TH. The use of porcupine inhibitors to target Wnt-driven cancers. Bioorg Med Chem Lett. 2015;25(23):5472–6. Epub 20151023. doi: 10.1016/j.bmcl.2015.10.032. PubMed PMID: 26522946.

60. Poulsen A, Ho SY, Wang W, Alam J, Jeyaraj DA, Ang SH, et al. Pharmacophore Model for Wnt/Porcupine Inhibitors and Its Use in Drug Design. J Chem Inf Model. 2015;55(7):1435–48. Epub 20150615. doi: 10.1021/acs.jcim.5b00159. PubMed PMID: 26024410.

61. Shah K, Panchal S, Patel B. Porcupine inhibitors: Novel and emerging anti-cancer therapeutics targeting the Wnt signaling pathway. Pharmacol Res. 2021;167:105532. Epub 20210304. doi: 10.1016/j.phrs.2021.105532. PubMed PMID: 33677106.

62. Flanagan DJ, Woodcock SA, Phillips C, Eagle C, Sansom OJ. Targeting ligand-dependent wnt pathway dysregulation in gastrointestinal cancers through porcupine inhibition. Pharmacol Ther. 2022;238:108179. Epub 20220328. doi: 10.1016/j.pharmthera.2022.108179. PubMed PMID: 35358569.

63. Rodon J, Argilés G, Connolly RM, Vaishampayan U, de Jonge M, Garralda E, et al. Phase 1 study of single-agent WNT974, a first-in-class Porcupine inhibitor, in patients with advanced solid tumours. British Journal of Cancer. 2021;125(1):28–37. doi: 10.1038/s41416-021-01389-8.

64. Janku F, de Vos F, de Miguel M, Forde P, Ribas A, Nagasaka M, et al. Abstract CT034: Phase I study of WNT974 + spartalizumab in patients (pts) with advanced solid tumors. Cancer Research. 2020;80(16_Supplement):CT034–CT. doi: 10.1158/1538-7445.am2020-ct034.

65. Jia XQ, Zhang S, Zhu HJ, Wang W, Zhu JH, Wang XD, et al. Increased Expression of PHGDH and Prognostic Significance in Colorectal Cancer. Transl Oncol. 2016;9(3):191–6. Epub 20160510. doi: 10.1016/j.tranon.2016.03.006. PubMed PMID: 27267836; PubMed Central PMCID: PMCPMC4907894.

66. Qian C, Xia Y, Ren Y, Yin Y, Deng A. Identification and validation of PSAT1 as a potential prognostic factor for predicting clinical outcomes in patients with colorectal carcinoma. Oncol Lett. 2017;14(6):8014–20. Epub 20171018. doi: 10.3892/ol.2017.7211. PubMed PMID: 29344244; PubMed Central PMCID: PMCPMC5755227.

67. Zhang J, Zhu Z, Miao Z, Huang X, Sun Z, Xu H, et al. The Clinical Significance and Mechanisms of REG4 in Human Cancers. Front Oncol. 2020;10:559230. Epub 20210108. doi: 10.3389/fonc.2020.559230. PubMed PMID: 33489872; PubMed Central PMCID: PMCPMC7819868.

68. Zheng HC, Xue H, Zhang CY. REG4 promotes the proliferation and anti-apoptosis of cancer. Front Cell Dev Biol. 2022;10:1012193. Epub 20220912. doi: 10.3389/fcell.2022.1012193. PubMed PMID: 36172286; PubMed Central PMCID: PMCPMC9511136.

69. Chou CL, Chen TJ, Tian YF, Chan TC, Yeh CF, Li WS, et al. CTSE Overexpression Is an Adverse Prognostic Factor for Survival among Rectal Cancer Patients Receiving CCRT. Life (Basel). 2021;11(7). Epub 20210702. doi: 10.3390/life11070646. PubMed PMID: 34357018; PubMed Central PMCID: PMCPMC8304221.

70. Kuester D, Lippert H, Roessner A, Krueger S. The cathepsin family and their role in colorectal cancer. Pathol Res Pract. 2008;204(7):491–500. Epub 20080624. doi: 10.1016/j.prp.2008.04.010. PubMed PMID: 18573619.

71. Sninsky JA, Bishnupuri KS, González I, Trikalinos NA, Chen L, Dieckgraefe BK. Reg4 and its downstream transcriptional activator CD44ICD in stage II and III colorectal cancer. Oncotarget. 2021;12(4):278–91. Epub 20210216. doi: 10.18632/oncotarget.27896. PubMed PMID: 33659040; PubMed Central PMCID: PMCPMC7899555.

72. Le CC, Bennasroune A, Langlois B, Salesse S, Boulagnon-Rombi C, Morjani H, et al. Functional Interplay Between Collagen Network and Cell Behavior Within Tumor Microenvironment in Colorectal Cancer. Front Oncol. 2020;10:527. Epub 20200430. doi: 10.3389/fonc.2020.00527. PubMed PMID: 32426274; PubMed Central PMCID: PMCPMC7204546.

73. Zhang Z, Wang Y, Zhang J, Zhong J, Yang R. COL1A1 promotes metastasis in colorectal cancer by regulating the WNT/PCP pathway. Mol Med Rep. 2018;17(4):5037–42. Epub 20180201. doi: 10.3892/mmr.2018.8533. PubMed PMID: 29393423; PubMed Central PMCID: PMCPMC5865965.

74. Zou X, Feng B, Dong T, Yan G, Tan B, Shen H, et al. Up-regulation of type I collagen during tumorigenesis of colorectal cancer revealed by quantitative proteomic analysis. J Proteomics. 2013;94:473–85. Epub 20131024. doi: 10.1016/j.jprot.2013.10.020. PubMed PMID: 24332065.

75. Xu S, Xu H, Wang W, Li S, Li H, Li T, et al. The role of collagen in cancer: from bench to bedside. J Transl Med. 2019;17(1):309. Epub 20190914. doi: 10.1186/s12967-019-2058-1. PubMed PMID: 31521169; PubMed Central PMCID: PMCPMC6744664.

76. Boland PM, Ma WW. Immunotherapy for Colorectal Cancer. Cancers (Basel). 2017;9(5). Epub 20170511. doi: 10.3390/cancers9050050. PubMed PMID: 28492495; PubMed Central PMCID: PMCPMC5447960.

77. Fridman WH, Pagès F, Sautès-Fridman C, Galon J. The immune contexture in human tumours: impact on clinical outcome. Nat Rev Cancer. 12. England 2012. p. 298–306.

78. Becht E, de Reyniès A, Giraldo NA, Pilati C, Buttard B, Lacroix L, et al. Immune and Stromal Classification of Colorectal Cancer Is Associated with Molecular Subtypes and Relevant for Precision Immunotherapy. Clin Cancer Res. 2016;22(16):4057–66. Epub 20160318. doi: 10.1158/1078-0432.ccr-15-2879. PubMed PMID: 26994146.

