## Supplementary material for "A novel molecular analysis approach in colorectal cancer suggests new treatment opportunities": sup figures

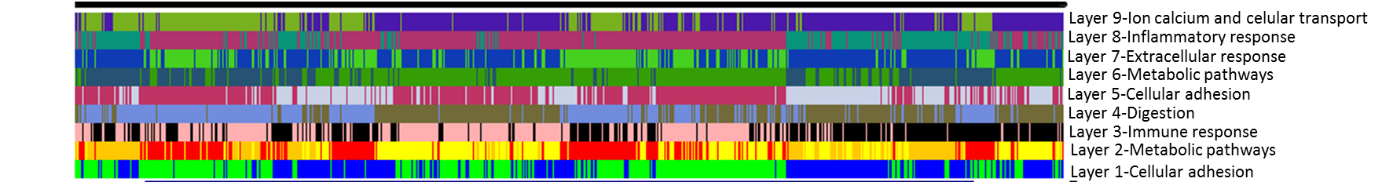

Sup Figure 1: Classifications obtained from biological layer analysis.

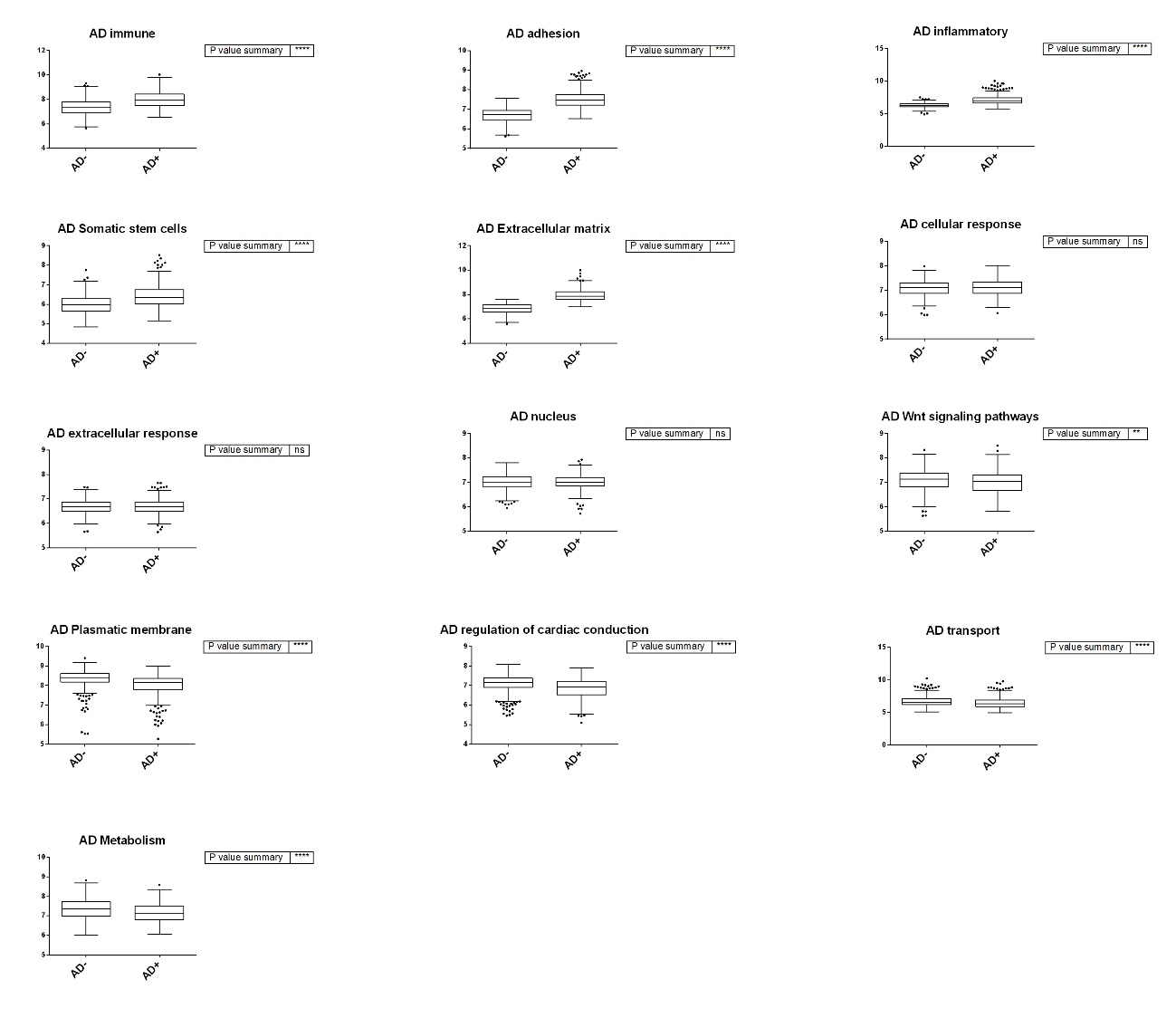

Sup Figure 2: Boxplots representing the activity of each functional node for the adhesion groups. High adhesion group (AD+), low adhesion group (AD-). (*p≤0.05, ** p≤ 0.01, *** p≤0.001, **** p≤0.0001); ns: non significance (p>0.05).

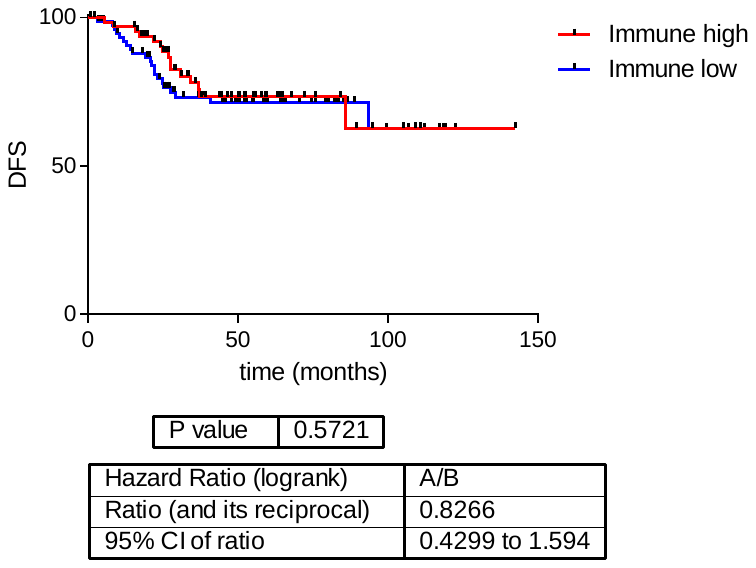

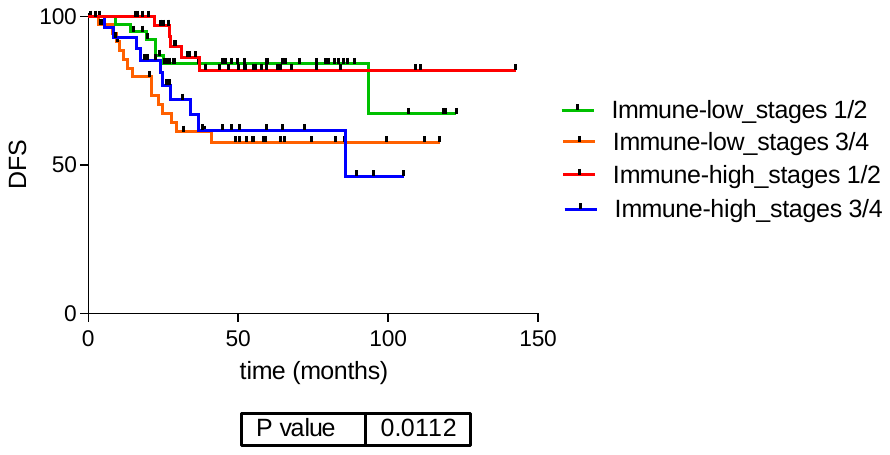

Sup Fig 3: Survival analysis of the immune groups. Immune high and immune low tumor prognosis was not significantly different. DFS: disease free survival.

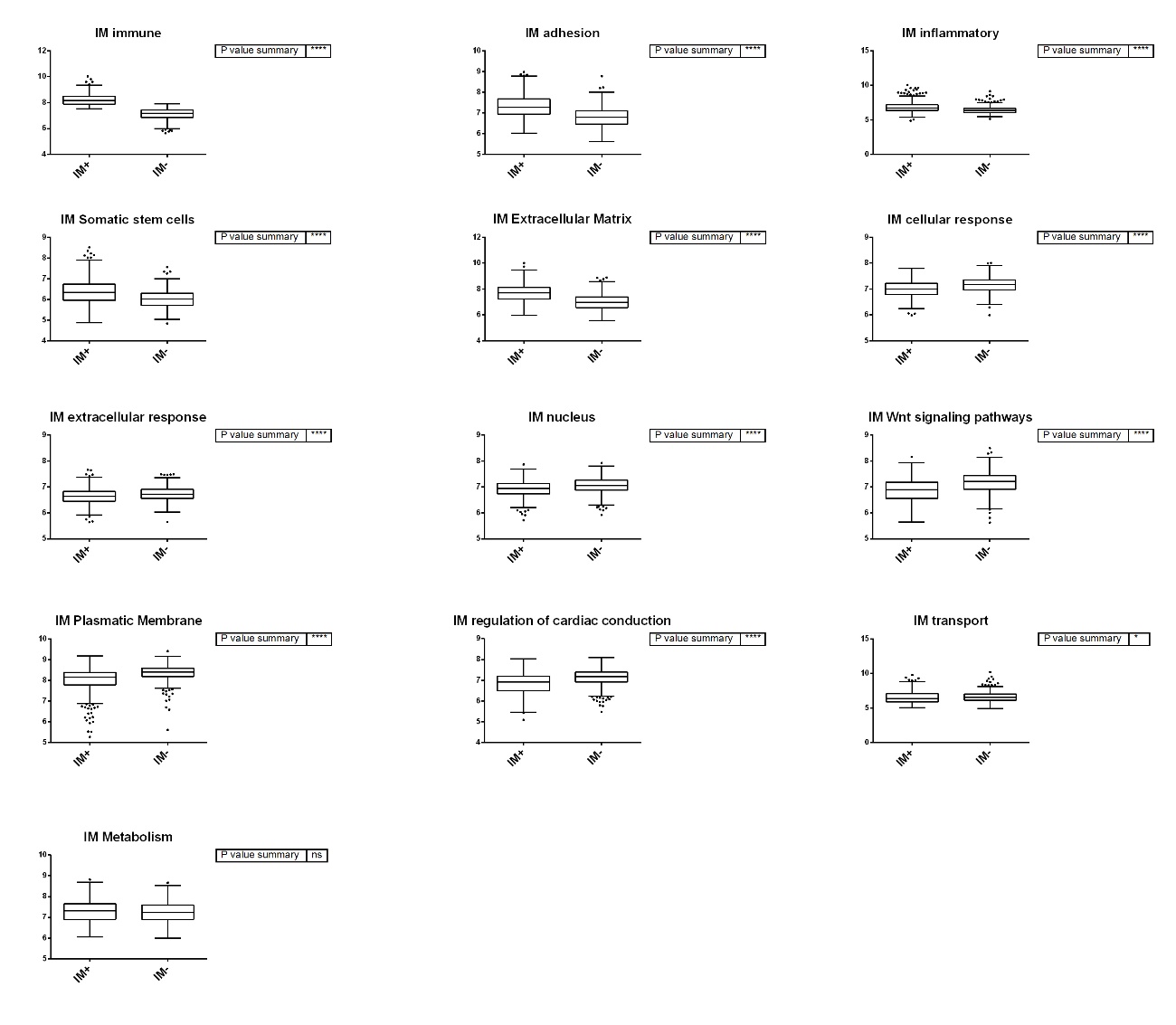
**Sup Figure 4:** Boxplots representing the activity of each functional node for the immune groups. Immune high group (IM+), immune low group (IM-). (*p≤0.05, ** p≤ 0.01, *** p≤0.001, **** p≤0.0001); ns: non significance (P>0.05).

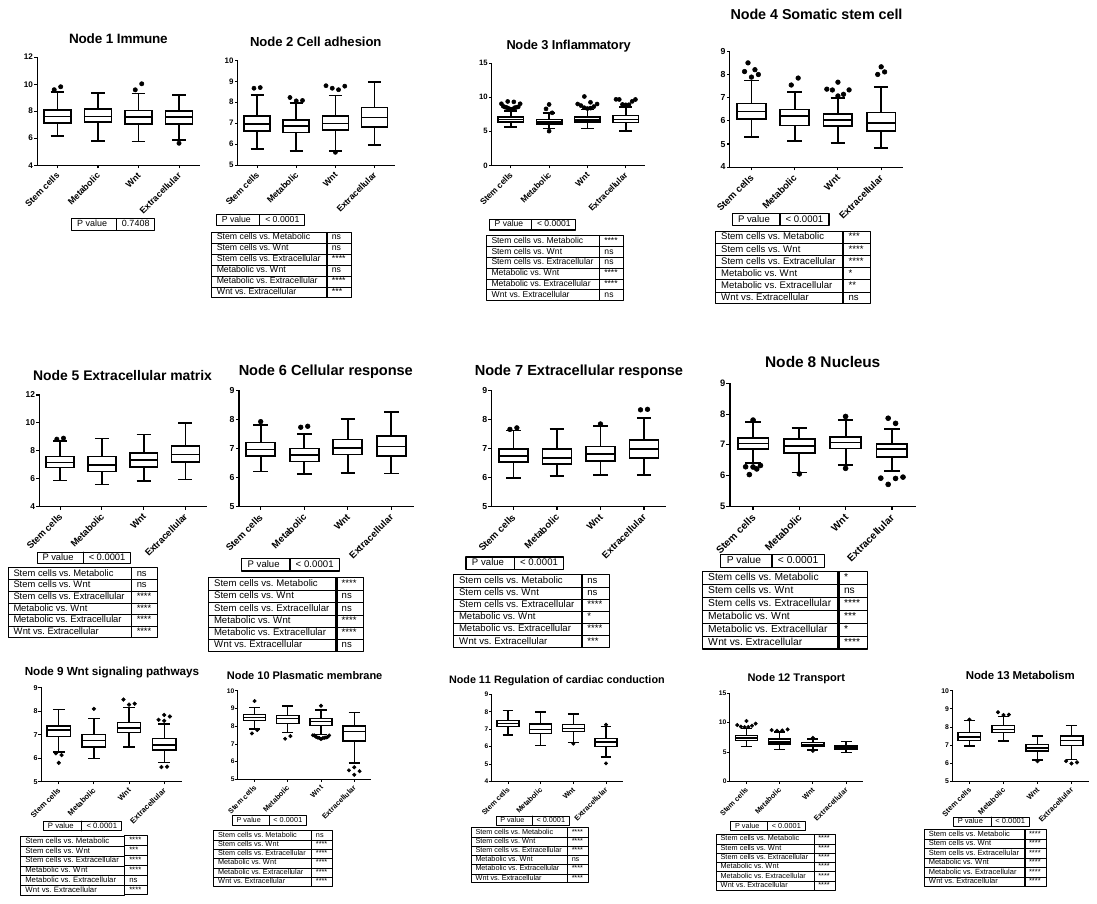

**Sup Figure 5**: Boxplots representing the activity of each functional node of the four molecular cluster subgroups (M1, M2, M3 and M4). (*p≤0.05, ** p≤0.01, *** p≤0.001, **** p≤0.0001); ns: non significance (p>0.05).

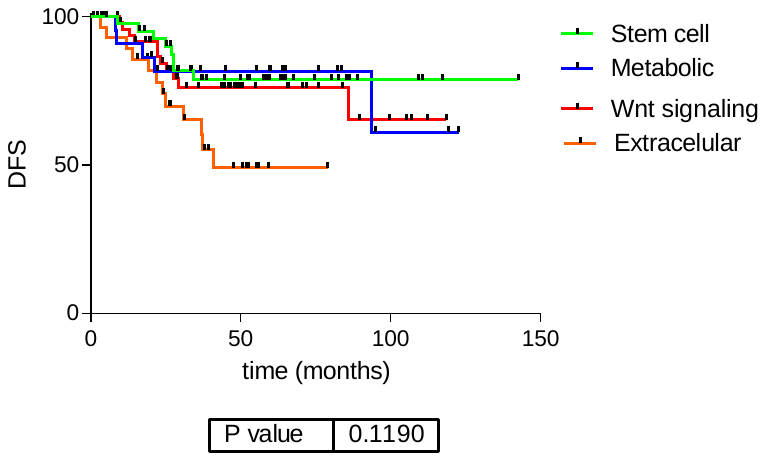

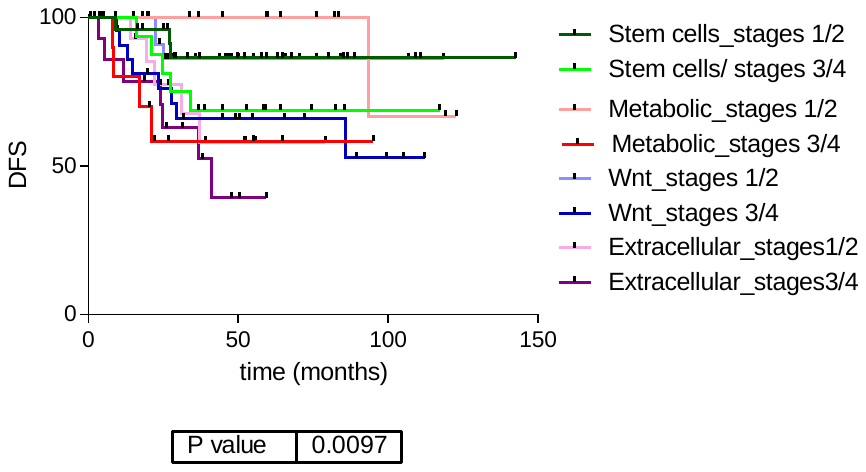

Sup Figure 6: Survival analyses of the four molecular groups. ns: non significance. DFS: disease free survival.
