## Supplementary material for "A novel molecular analysis approach in colorectal cancer suggests new treatment opportunities": sup tables

| Number of patients | 805 (100%) |
| --- | --- |
| Age median (range) | 66 (26-92) |
| Gender |  |
| Male | 96 (12%) |
| Female | 81 (10%) |
| Unknown | 628 (78%) |
| Location |  |
| Distal | 342 (42%) |
| Proximal | 224 (28%) |
| Unknown | 239 (30%) |
| 8th ed. TNM AJCC Stage |  |
| 1 | 57 (7%) |
| 2 | 321 (40%) |
| 3 | 262 (33%) |
| 4 | 99 (12%) |
| Unknown | 66 (8%) |

Sup Table 1: Clinical characteristics of CRC patients.

|  |  | | CMS1 | CMS2 | CMS3 | CMS4 | NOLBL |
| --- | --- | --- | --- | --- | --- | --- | --- |
| ADHESION | **low adhesion** | | 68 (52%) | 255 (81%) | 95 (98%) | 2 (2%) | 34 (47%) |
|  | **high adhesion** | | 64 (48%) | 60 (19%) | 2 (2%) | 186 (99%) | 39 (53%) |
| IMMUNE | **immune positive** | | 106 (80%) | 57 (18%) | 18 (19%) | 139 (74%) | 44 (60%) |
|  | **immune negative** | | 26 (20%) | 258 (82%) | 79 (81%) | 49 (26%) | 29 (40%) |
| MOLECULAR | **Stem cells** | | 2 (2%) | 123 (39%) | 18 (19%) | 42 (22%) | 36 (49%) |
|  | **Metabolic** | | 38 (29%) | 0 (0%) | 73 (75%) | 12 (6%) | 14 (19%) |
|  | **Wnt pathway** | | 3 (2%) | 191 (60.7%) | 0 (0%) | 88 (47%) | 18 (25%) |
|  | **Extracellular** | | 89 (67%) | 1 (0.3%) | 6 (6%) | 46 (24%) | 5 (7%) |

Sup Table 5: Number of patients and percentage of each consensus molecular subtype (CMS) assigned to immune, adhesion, and molecular layers.
